# Skeletal muscle energetics explain the sex disparity in mobility impairment in the Study of Muscle, Mobility and Aging (SOMMA)

**DOI:** 10.1101/2023.11.08.23298271

**Authors:** Philip A. Kramer, Paul M. Coen, Peggy M. Cawthon, Giovanna Distefano, Steven R. Cummings, Bret H. Goodpaster, Russell T. Hepple, Stephen B. Kritchevsky, Eric G. Shankland, David J. Marcinek, Frederico G. S. Toledo, Kate A. Duchowny, Sofhia V. Ramos, Stephanie Harrison, Anne B. Newman, Anthony J. A. Molina

**Author notes:** Corresponding Authors: Anthony J. A. Molina, PhD, University of California San Diego School of Medicine, 9500 Gilman Drive, La Jolla, CA. 92093, Philip Kramer, PhD, Wake Forest University School of Medicine 1 Medical Center Blvd., Winston-Salem, NC 27157.

## Abstract

The age-related decline in muscle mitochondrial energetics contributes to the loss of mobility in older adults. Women experience a higher prevalence of mobility impairment compared to men, but it is unknown whether sex-specific differences in muscle energetics underlie this disparity. In the Study of Muscle, Mobility and Aging (SOMMA), muscle energetics were characterized using in vivo phosphorus-31 magnetic resonance spectroscopy and high-resolution respirometry of vastus lateralis biopsies in 773 participants (56.4% women, age 70-94 years). A Short Physical Performance Battery score ≤ 8 was used to define lower-extremity mobility impairment. Muscle mitochondrial energetics were lower in women compared to men (e.g. Maximal Complex I&II OXPHOS: Women=55.06 +/− 15.95; Men=65.80 +/− 19.74; p<0.001) and in individuals with mobility impairment compared to those without (e.g., Maximal Complex I&II OXPHOS in women: SPPB≥9=56.59 +/− 16.22; SPPB≤8=47.37 +/− 11.85; p<0.001). Muscle energetics were negatively associated with age only in men (e.g., Maximal ETS capacity: R=-0.15, p=0.02; age/sex interaction, p=0.04), resulting in muscle energetics measures that were significantly lower in women than men in the 70-79 age group but not the 80+ age group. Similarly, the odds of mobility impairment were greater in women than men only in the 70-79 age group (70-79 age group, OR_age-adjusted_=1.78, 95% CI=1.03, 3.08, p=0.038; 80+ age group, OR_age-adjusted_=1.05, 95% CI=0.52, 2.15, p=0.89). Accounting for muscle energetics attenuated up to 75% of the greater odds of mobility impairment in women. Women had lower muscle mitochondrial energetics compared to men, which largely explain their greater odds of lower-extremity mobility impairment.

## Introduction

The age-related decline of mitochondrial oxidative capacity in skeletal muscle is believed to contribute to mobility impairment, which affect the independence, health, and finances of nearly a third of all men and women over the age of 65 in the US ^1–7^. Evidence suggests mobility impairment and decreased physical activity increases the risk of falls, hospitalizations, and chronic health conditions such as diabetes and dementia ^8–10^. Mobility impairment disproportionately affects women, who are more likely to develop mobility impairment and at a younger age than men ^11–14^. The underlying etiology for the sex disparities in mobility impairment is partly explained by a higher prevalence of arthritis and greater adiposity in women, but differences in muscle energetics have not been examined^15^. Several studies have reported a difference in mitochondrial energetics in the muscle of men and women, citing differences in mitochondrial protein and cardiolipin content, ADP and calcium sensitivity, fatty acid oxidation, oxidative stress, and the estrogen response ^16–19^. However, no studies have addressed sex differences in muscle energetics within an older population for its contribution to the observed sex disparity in mobility impairment.

High-resolution Respirometry (HHR) of permeabilized fibers from skeletal muscle biopsies has been utilized in numerous studies to assess mitochondrial respiratory capacity including the specific contributions of mitochondrial complexes ^20^. Mitochondrial respiration in muscle has been shown to correlate with age, physical function, and clinical outcomes associated with sarcopenia ^1,5,21–25^. Phosphorus-31 magnetic resonance spectroscopy (^31^P MRS), by contrast, is non-invasive and can measure mitochondrial oxidative capacity indirectly through modeling phosphocreatine recovery following a brief isometric kicking exercise ^20^. The Baltimore Longitudinal Study of Aging demonstrated the value of ^31^P MRS assessment of mitochondrial oxidative capacity as a predictor of gait speed and mobility decline, while other studies have shown the effect of age, physical function, and exercise on muscle energetics ^2,4,24,26–28^. Together, in vivo ^31^P MRS and ex vivo HRR of permeabilized muscle fibers provide a complementary and comprehensive assessment of mitochondrial energetics that improve the association and predictive value of muscle metabolism on phenotypes relevant to aging, including gait speed and aerobic capacity ^4^.

The Study of Muscle, Mobility and Aging (SOMMA) is a longitudinal, observational, multi-site cohort study with a focus on the biological basis of muscle aging and mobility decline ^29^. In this analysis of SOMMA baseline data, we sought to evaluate sex differences in skeletal muscle energetics using both in vivo ^31^P MRS and ex vivo permeabilized muscle respirometry and determine its role in lower-extremity mobility impairment. Here, we test the hypothesis that significant differences in muscle energetics exist between older men and women, and that lower muscle mitochondrial energetics help explain the greater prevalence of mobility impairment in women.

## Methods

### Study cohort and recruitment

The Study of Muscle, Mobility and Aging (SOMMA; https://www.sommastudy.com/) recruited and consented 879 participants aged 70 years and older at The University of Pittsburgh and Wake Forest University School of Medicine from April 2019 to December 2021 as approved by WIRB-Copernicus Group (WCG IRB)(20180764)^29^. SOMMA collected baseline physical function and muscle energetics data on 773 (337, 43.6% men; 436, 56.4% women) participants aged 70 to 94 at Wake Forest University and the University of Pittsburgh. Participants were eligible for this study if willing and able to undergo muscle tissue biopsy, magnetic resonance spectroscopy (MRS), walk 400 meters or climb a flight of stairs, and had no medical contraindications to procedures, no active malignancy or dementia, and had a walking speed of ≥ 0.6m/s and BMI <40kg/m^2^. For this study, self-reported sex was used for group assignment.

### Health and Physical function assessments

SOMMA baseline health and physical function assessments included a Short Physical Performance Battery (SPPB), BMI, and self-reported health history and number of chronic health conditions (multimorbidity), medications, diet, and physical activity as measured as weekly caloric expenditure from activities on the Community Healthy Activities Model Program for Seniors (CHAMPS) questionnaire as previously described ^24^. Total activity count, an empirical measure of physical activity, was obtained from a wrist-worn actigraphy device which collected accelerometry data on all 3 orthogonal axes (ActiGraph GTX9). The device was worn continuously for 7 days at baseline and total activity counts were reported as the mean of the 24-hr total (imputed). Study staff performed the SPPB as developed and described by the National Institute on Aging (https://sppbguide.com/), and scored participants on a scale from 0-12 (total), or the sum of a balance test, walking speed test, and chair standing test ^30^. Chair stands were analyzed as the number of chair stands per ten seconds. Individuals unable to complete a chair stand in 10 seconds were given a value of 0 stands/10 seconds. 400m and 4m gait speed were analyzed as meters per second. Muscle mass was obtained by a D3-creatine dilution protocol and measured in the urine by high-performance liquid chromatography and tandem mass spectroscopy (MS/MS) as previously described ^29^.The number of chronic health conditions was assessed using the multimorbidity index based on Rochester Epidemiology Project (Range 0-13) to include history of cancer, excluding non-melanoma skin cancer, history of heart disease (congenital heart defects, congestive heart failure, atrial fibrillation, aortic stenosis), history of arthritis, chronic kidney disease, stroke, diabetes, dementia, depression, COPD and osteoporosis. Lower extremity mobility impairment was defined as having an SPPB score less than or equal to 8, a threshold previously used in the International Mobility in Aging Study and recommended by the European Working Group on Sarcopenia in Older People for the screening of poor physical performance and sarcopenia ^31–33^.

### Muscle biopsy and fiber bundle processing

Vastus lateralis biopsies were conducted at the clinical sites as previously described ^24^, and approximately 20mg of tissue was cleaned of blood, fat, and connective tissue before being placed in ice-cold BIOPS media (10mM Ca–EGTA buffer, 0.1M free calcium, 20mM imidazole, 20mM taurine, 50mM potassium 2-[N-morpholino]-ethanesulfonic acid, 0.5mM dithiothreitol, 6.56mM MgCl2, 5.77mM ATP, and 15mM phosphocreatine [PCr], pH 7.1). Within 2 hours, the muscle tissue was transported to the processing laboratory where it was dissected into pieces approximately 3mg in size, and the fiber bundles teased apart while submerged in ice-cold BIOPS. Permeabilization of the prepared fiber bundles took place in 2ml of BIOPS containing 50ug/mL saponin for each bundle. Tubes were rocked for 30 min on ice. Ice-cold MiR05 media (0.5mM EDTA, 3mM MgCl2·6H2O, 60mM K-lactobionate, 20mM taurine, 10mM KH2PO4, 20mM HEPES, 110mM sucrose, and 1g/L BSA, pH 7.1) was used to wash the fiber bundles twice for ten minutes each, with blebbistatin (25μM) added to the second wash. Fiber bundles were then placed on filter paper to absorb excess media for 5 seconds, turned and blotted for 2 seconds, and weighed to obtain the wet weight. Fiber bundle target weights were 2-3mgs.

### High resolution respirometry (HRR) protocol

Following air calibration with MiR05 supplemented with blebbistatin (25μM) at 37°C, permeabilized vastus lateralis fiber bundles were immediately placed in the Oroboros O2k Oxygraph (Oroboros Inc., Innsbruck, Austria) in duplicate as previously described ^24^. Briefly, oxygen levels were raised to 400μM just prior to beginning the substrate uncoupler inhibitor titration (SUIT) protocol. Complex I-supported leak respiration was measured following the addition of pyruvate (5mM) and malate (2mM), Complex I OXPHOS was obtained after the addition of ADP (4.2mM), Maximal Complex I supported OXPHOS after addition of glutamate (10mM), Maximal Complex I&II supported OXPHOS upon the addition of succinate (10mM), and Maximal ETS capacity after FCCP (1μM increments) was titrated to maximal oxygen consumption. All oxygen consumption measurements (pmol/s) were normalized to fiber bundle wet weight (pmol/s*mg). The Respiratory control ratio (RCR) was calculated as the ratio between Complex I supported OXPHOS and Complex I-supported leak respiration. Additional quality control measures included cytochrome c (10µM) injection to ensure mitochondrial outer membrane integrity. Samples with a greater than 15% change were omitted from the dataset. Approximately 6.6% of the fiber bundles analyzed showed an increase in respiration between 10-15% after injection of cytochrome c. The coefficient of variation for O2k chamber duplicates for maximal complex I&II-supported OXPHOS was 11% for Wake Forest and 12% for the University of Pittsburgh. All data from both clinical sites were analyzed in Datlab 7.4 software.

### In Vivo ^31^P MRS protocol

In vivo muscle mitochondrial energetics were measured in participant quadriceps using ^31^P MRS with a 3 Tesla MR magnet (Siemen’s Medical System—Prisma (University of Pittsburgh) or Skyra (Wake Forest University School of Medicine) as previously described ^24^. Briefly, the supine participant had their right knee joint positioned in 20°-30° of flexion with their right vastus lateralis muscle centered to a 12” 31P/1H dual-tuned, surface RF coil (PulseTeq, Limited). After training, the participant performed a bout of repeated isometric knee extension (∼30 sec) with resistance applied by an ankle strap. A second bout of kicking followed, adjusted for length of time (18-36 sec), to achieve adequate PCr breakdown while maintaining intracellular pH above 6.8. The rate of phosphocreatine (PCr) recovery following the acute bout of knee extensor exercise was used to calculate maximal mitochondrial ATP production (ATPmax) by dividing the time-constant of a monoexponential fit of PCr recovery by an assumed resting PCr value of 24.5mM. The mean coefficient of variation for duplicates of ATPmax measurement was 9.9% across both clinic sites.

### Statistical Analyses

Differences in characteristics by sex were determined by using t-tests for continuous variables and chi-square tests for categorical variables. A Kruskal-Wallis test was used for skewed data. Relationships between age and muscle energetics variables by sex were assessed by Pearson correlations. T-tests were used to test for differences in muscle energetics between men and women or mobility impairment status and within each age group (70-74, 75-79, and 80+). Sex stratified linear regression was used to compare adjusted means and 95% confidence intervals for muscle energetics. We employed sex stratified, logistic regression to model the odds of having a SPPB score of less than or equal to 8. To assess mediation by mitochondrial energetics between the relationship of mobility impairment and sex, mitochondrial variables were added to the logistic models and the change in men and women due to this addition was evaluated within each age group (70-79, 80+). Similarly, the change in mobility impairment odds in men and women per 1 standard deviation increase in muscle energetics were also assessed. All models were adjusted for age, BMI, gait speed, race, technician, CHAMPS, and number of chronic conditions, unless otherwise specified. In logistic models with mobility impairment as the outcome, gait speed (a component of the SPPB) was excluded as a covariate.

## Results

### Sex Differences in Participant Characteristics

The Study of Muscle, Mobility and Aging performed in vivo and ex vivo muscle mitochondrial energetics assessments and measured the physical function of 337 men and 436 women from 2019 to early 2022. There was no difference in the age and race (% Non-Hispanic white) between men and women, however, men had a slightly higher BMI (p=0.0429) and fewer chronic health conditions (p=0.0004) compared to women (**Table 1**). Men also reported more caloric expenditure from physical activities as measured by CHAMPS (p<0.0001). Interestingly, women had higher total activity levels compared to men as measured by actigraphy, but had significantly lower muscle mass as determined by urine D3 creatinine. When analyzed as an ordinal outcome, SPPB was not significantly different between men and women (p=0.4387). However, 4 meter gait speed, a component of the SPPB, and 400 meter walk gait speed, when analyzed as meters per second, were significantly higher in men than women (p=0.0054 and p<0.0001, respectively). Chair stand performance did not differ between men and women. Defining mobility impairment as an SPPB score ≤ 8, women had a higher prevalence than men, though this was not significant (p=0.0751). This was driven by the 70-79 age group, with mobility impairment prevalence being significantly higher in women than men (p=0.0278), while the prevalence was approximately 26% in both men and women after the age of 80 (p=0.9910).

**Table 1.**
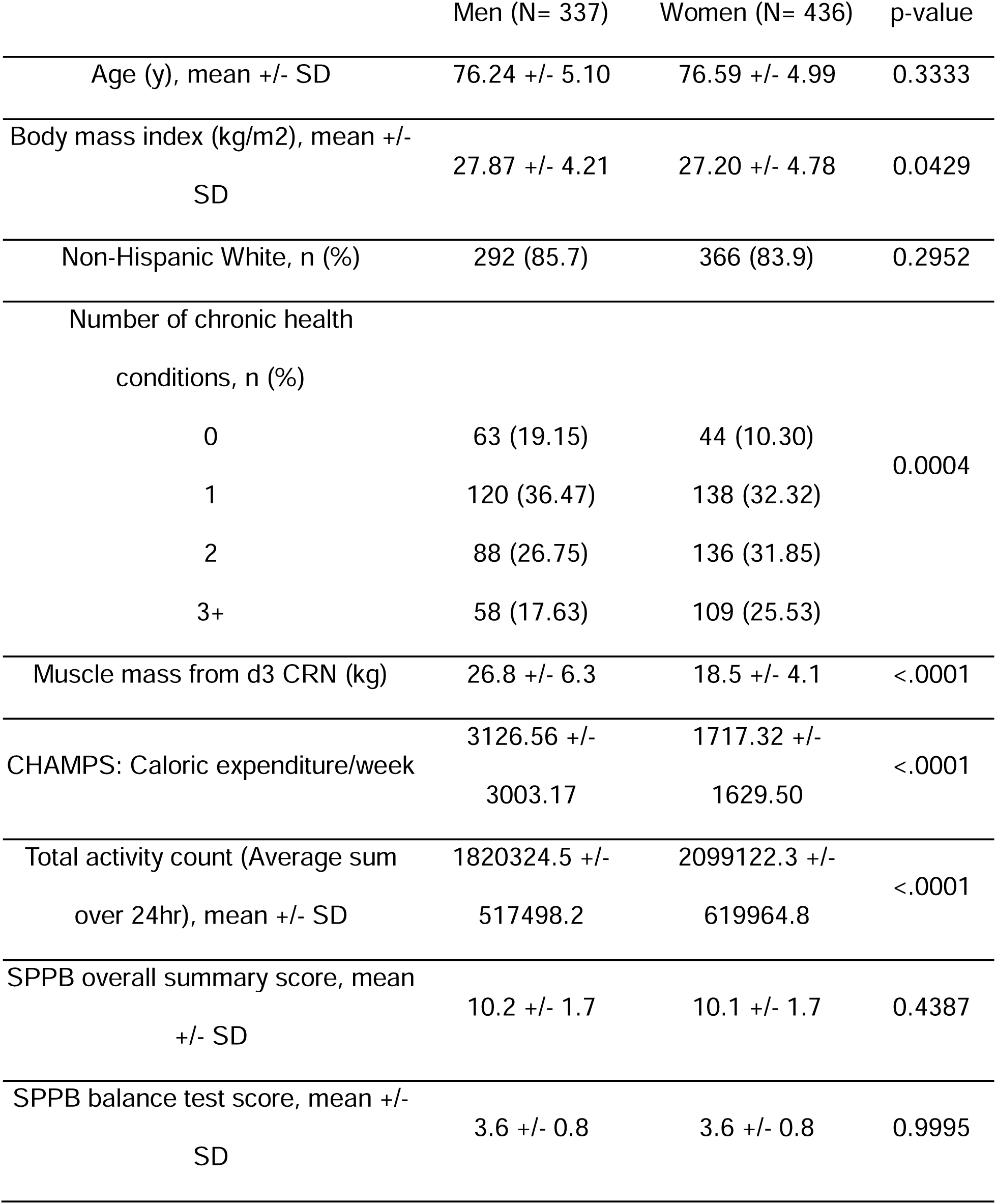
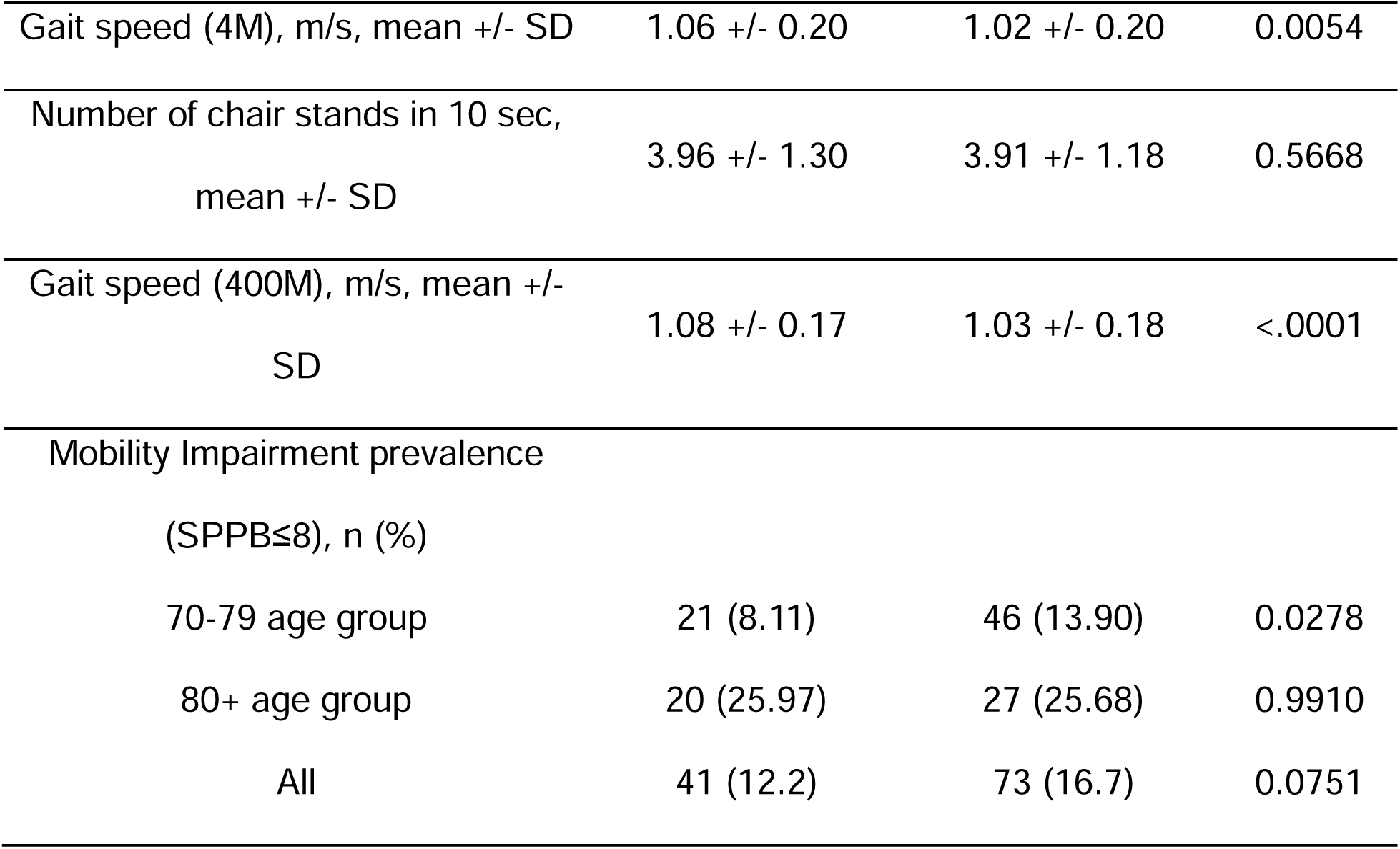
Characteristics of covariates by sex.

### Muscle Energetics Differences by Sex and Mobility Impairment

Muscle energetics were significantly greater in men across every in vivo and ex vivo measure (**Table 2**) but RCR and PCr/ATP ratio, the latter of which was significantly higher in women. To determine if any covariates explained sex differences in muscle energetics, we adjusted for age, BMI, gait speed, race, technician, CHAMPS, and number of chronic health conditions (**Table 2**). All muscle energetics measures, excluding RCR, remained significantly different between men and women, with the exception of ATPmax, which was no longer significantly different (p=0.3578).

**Table 2.**
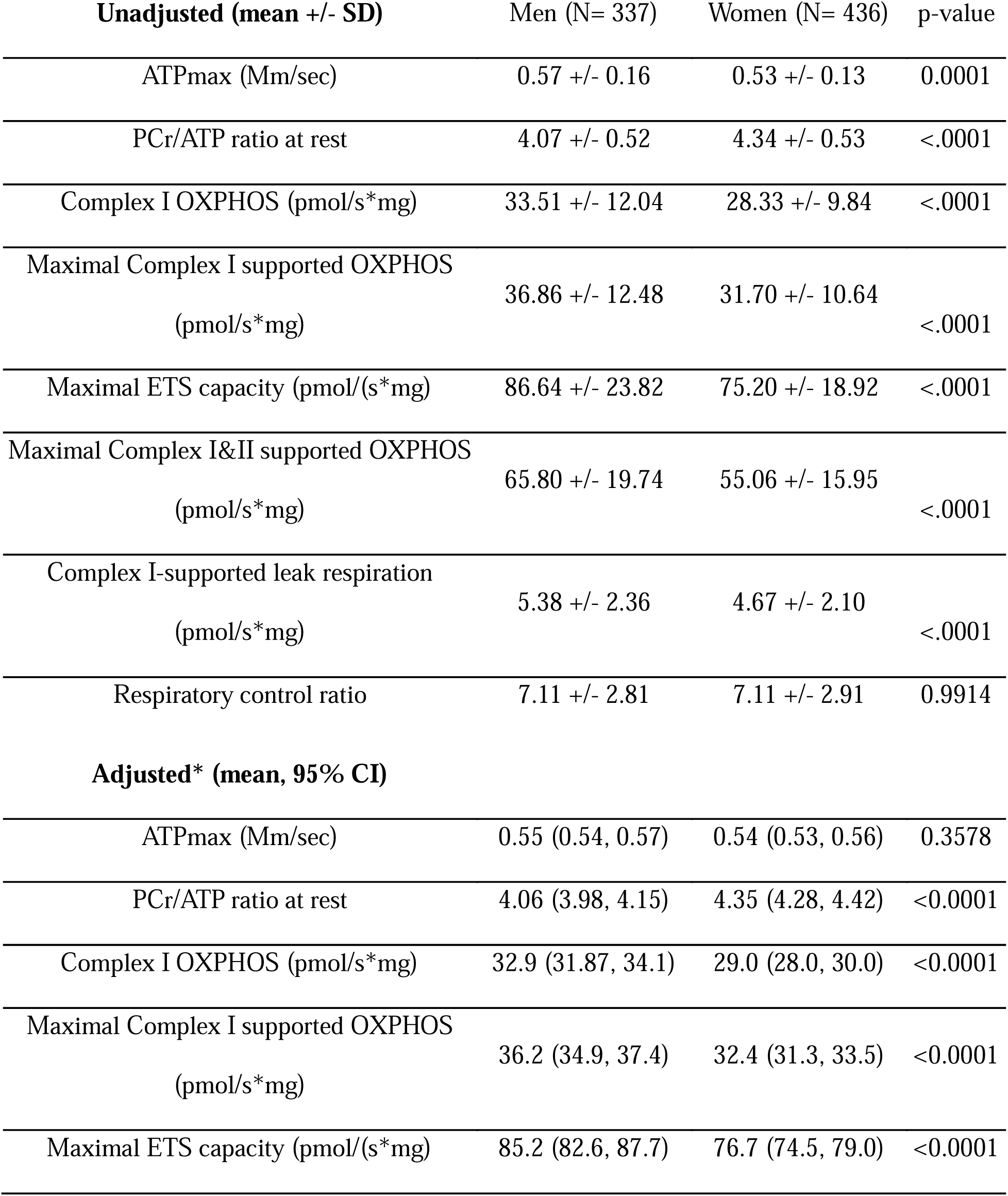
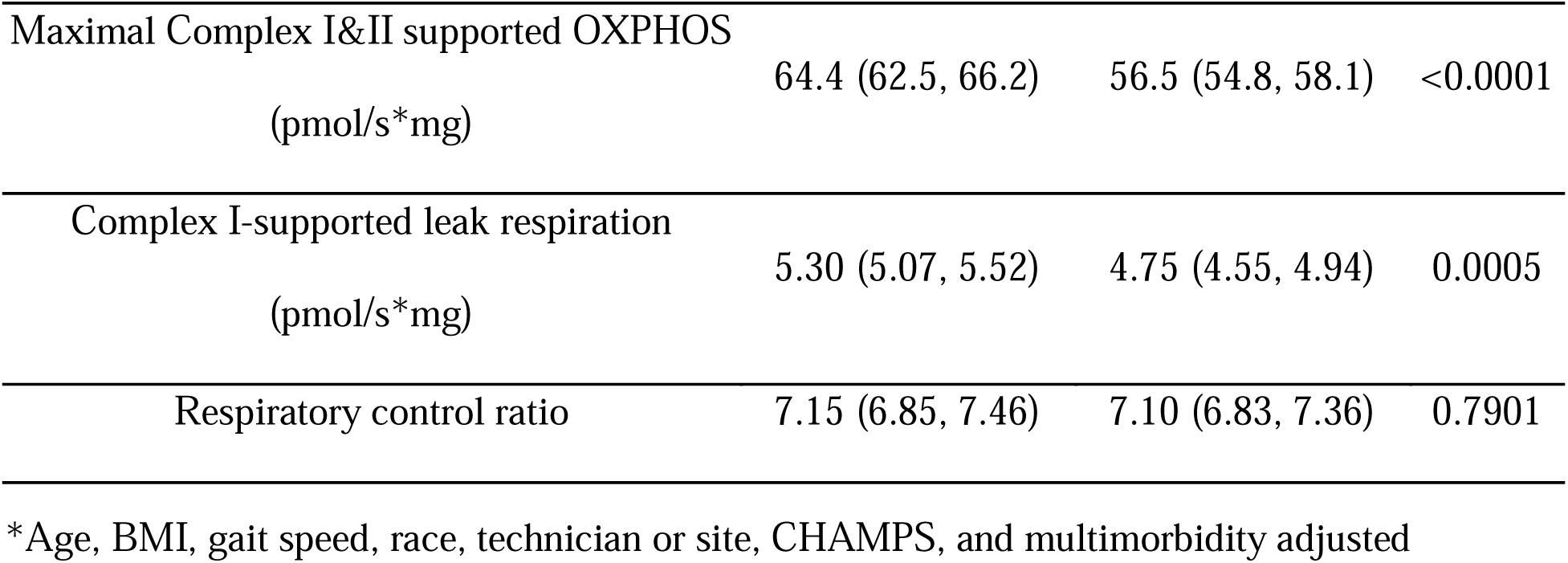
Unadjusted and Age, BMI, gait speed, race, technician or site, CHAMPS, and number of chronic health conditions adjusted sex differences in muscle energetics.

| **Unadjusted (mean +/- SD)** | **Men (N= 337)** | **Women (N= 436)** | **p-value** |
| --- | --- | --- | --- |
| ATPmax (Mm/sec) | 0.57 +/- 0.16 | 0.53 +/- 0.13 | 0.0001 |
| PCr/ATP ratio at rest | 4.07 +/- 0.52 | 4.34 +/- 0.53 | <.0001 |
| Complex I OXPHOS (pmol/s*mg) | 33.51 +/- 12.04 | 28.33 +/- 9.84 | <.0001 |
| Maximal Complex I supported OXPHOS<br>(pmol/s*mg) | 36.86 +/- 12.48 | 31.70 +/- 10.64 | <.0001 |
| Maximal ETS capacity (pmol/(s*mg)) | 86.64 +/- 23.82 | 75.20 +/- 18.92 | <.0001 |
| Maximal Complex I&II supported OXPHOS<br>(pmol/s*mg) | 65.80 +/- 19.74 | 55.06 +/- 15.95 | <.0001 |
| Complex I-supported leak respiration<br>(pmol/s*mg) | 5.38 +/- 2.36 | 4.67 +/- 2.10 | <.0001 |
| Respiratory control ratio | 7.11 +/- 2.81 | 7.11 +/- 2.91 | 0.9914 |
| **Adjusted\* (mean, 95% CI)** |  |  |  |
| ATPmax (Mm/sec) | 0.55 (0.54, 0.57) | 0.54 (0.53, 0.56) | 0.3578 |
| PCr/ATP ratio at rest | 4.06 (3.98, 4.15) | 4.35 (4.28, 4.42) | <0.0001 |
| Complex I OXPHOS (pmol/s*mg) | 32.9 (31.87, 34.1) | 29.0 (28.0, 30.0) | <0.0001 |
| Maximal Complex I supported OXPHOS<br>(pmol/s*mg) | 36.2 (34.9, 37.4) | 32.4 (31.3, 33.5) | <0.0001 |
| Maximal ETS capacity (pmol/(s*mg)) | 85.2 (82.6, 87.7) | 76.7 (74.5, 79.0) | <0.0001 |
| Maximal Complex I&II supported OXPHOS<br>(pmol/s*mg) | 64.4 (62.5, 66.2) | 56.5 (54.8, 58.1) | <0.0001 |
| Complex I-supported leak respiration<br>(pmol/s*mg) | 5.30 (5.07, 5.52) | 4.75 (4.55, 4.94) | 0.0005 |
| Respiratory control ratio | 7.15 (6.85, 7.46) | 7.10 (6.83, 7.36) | 0.7901 |

\*Age, BMI, gait speed, race, technician or site, CHAMPS, and multimorbidity adjusted

The relationship between muscle energetics and age was illustrated using age groups of 70-74, 75-79, 80+ years in both men and women (**Figure 1**). Interestingly, muscle energetics in women had no association with age while most muscle energetics measures in men were consistently negatively correlated with age except PCr/ATP ratio (**Figure 1B**) and RCR (**Figure 1H**). Of all the muscle energetics measures, Complex I-supported leak respiration (p=0.0304), Maximal Complex I&II supported OXPHOS (p=0.0892), Maximal ETS capacity (p=0.0422), Complex I OXPHOS (p=0.0438), and ATPmax (p=0.095) were significant or trended toward significance (p<0.10) for the age/sex interaction. As illustrated by age-range bound tertiles, all muscle energetics measures other than RCR were significantly different in men compared to women between the ages of 70-79, except ATPmax in the 75-79 age group (p=0.06). After 80 years of age, no sex differences were observed in muscle energetics with the exception of PCr/ATP ratio at rest (**Figure 1B**) and Maximal Complex I&II OXPHOS (**Figure 1E**), which still remained significant.

**Figure 1.**
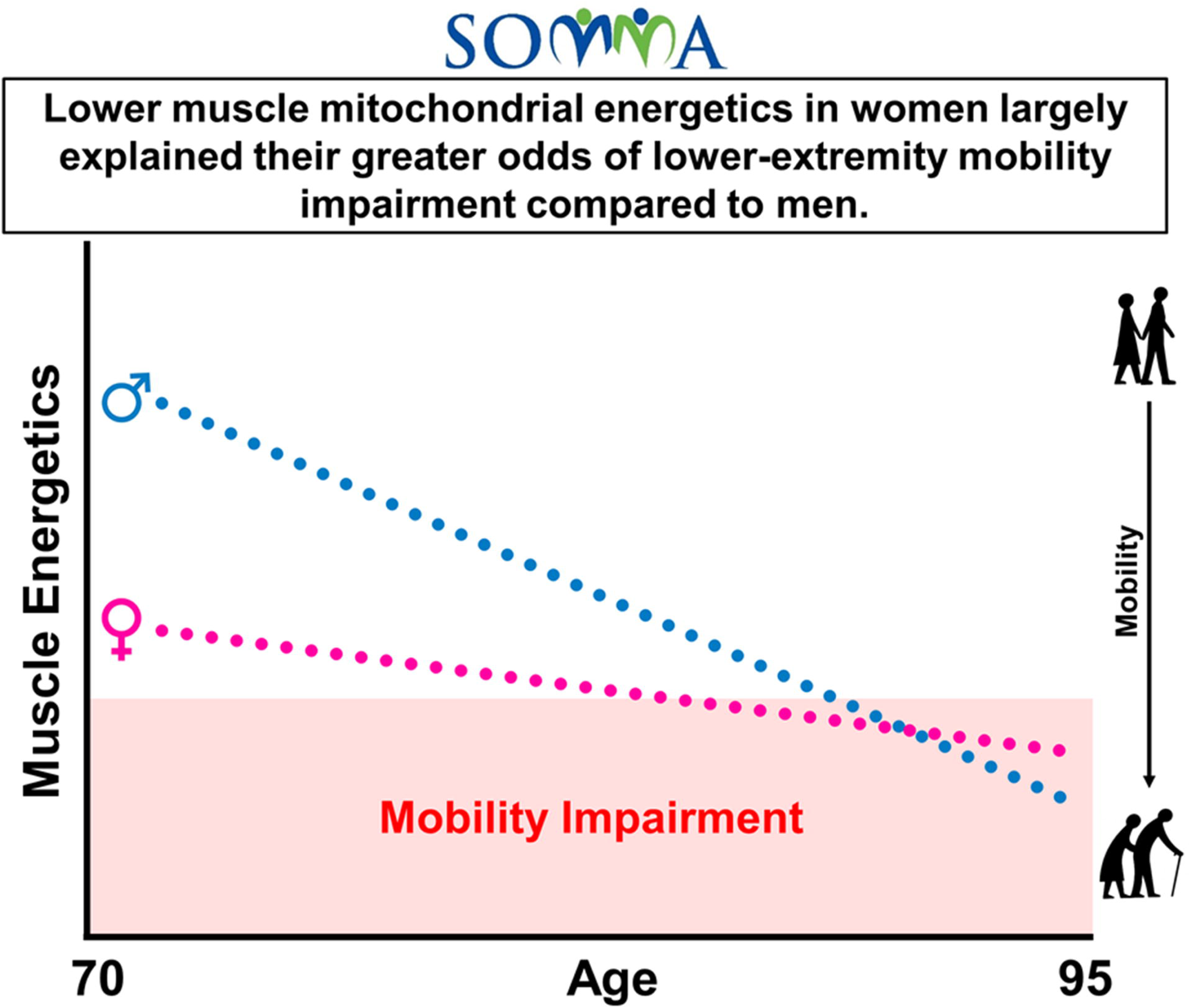
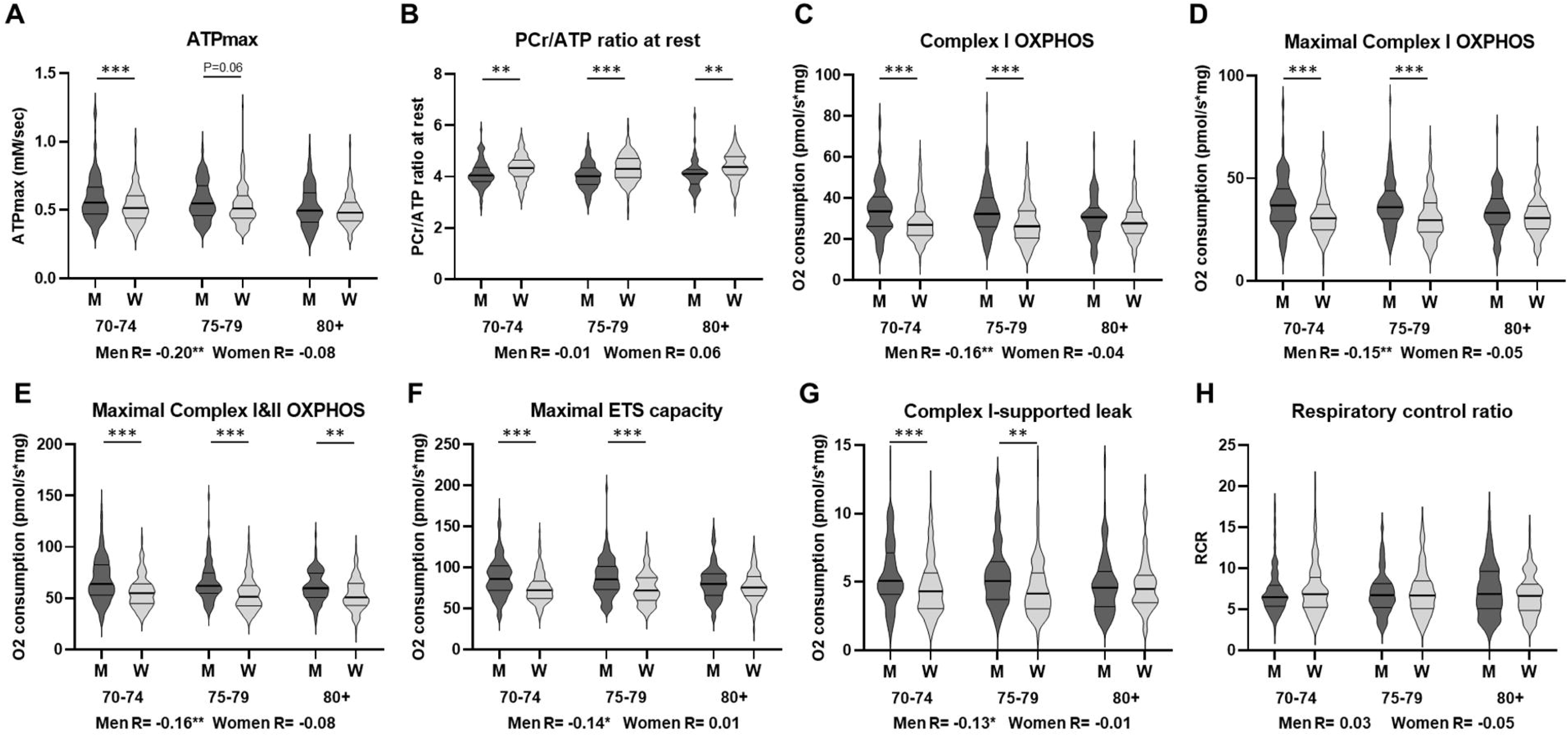
Sex differences in muscle energetics by age range-bound tertiles. In vivo **(A-B)** and ex vivo **(C-H)** muscle energetics measures (unadjusted) are shown by violin plot in age-range bound tertiles (70-74, 75-79, 80+) among men (M)(n=158, 101, 78) and women (W)(n=192, 139, 105). ***=p<0.001, **=p<0.01.

Mobility impairment status was associated with significantly lower Complex I OXPHOS, Maximal Complex I OXPHOS, and Maximal Complex I and II OXPHOS in both men and women (**Figure 2A**). Women with mobility impairment also had significantly lower ATPmax and Maximal ETS capacity. Interestingly, men with mobility impairment had comparable muscle energetics to women without mobility impairment. Due to the observed effects of age and other covariates on muscle energetics, especially in men, we further examined the adjusted means (without gait speed) by gender and mobility impairment status (**Figure 2B**). Following adjustment, there were no longer any significant differences between men with mobility impairment and men without mobility impairment. Women, however, still retained all significant differences in muscle energetics based on their mobility impairment status with the exception of ATPmax and Complex I OXPHOS (p=0.058).

**Figure 2:**
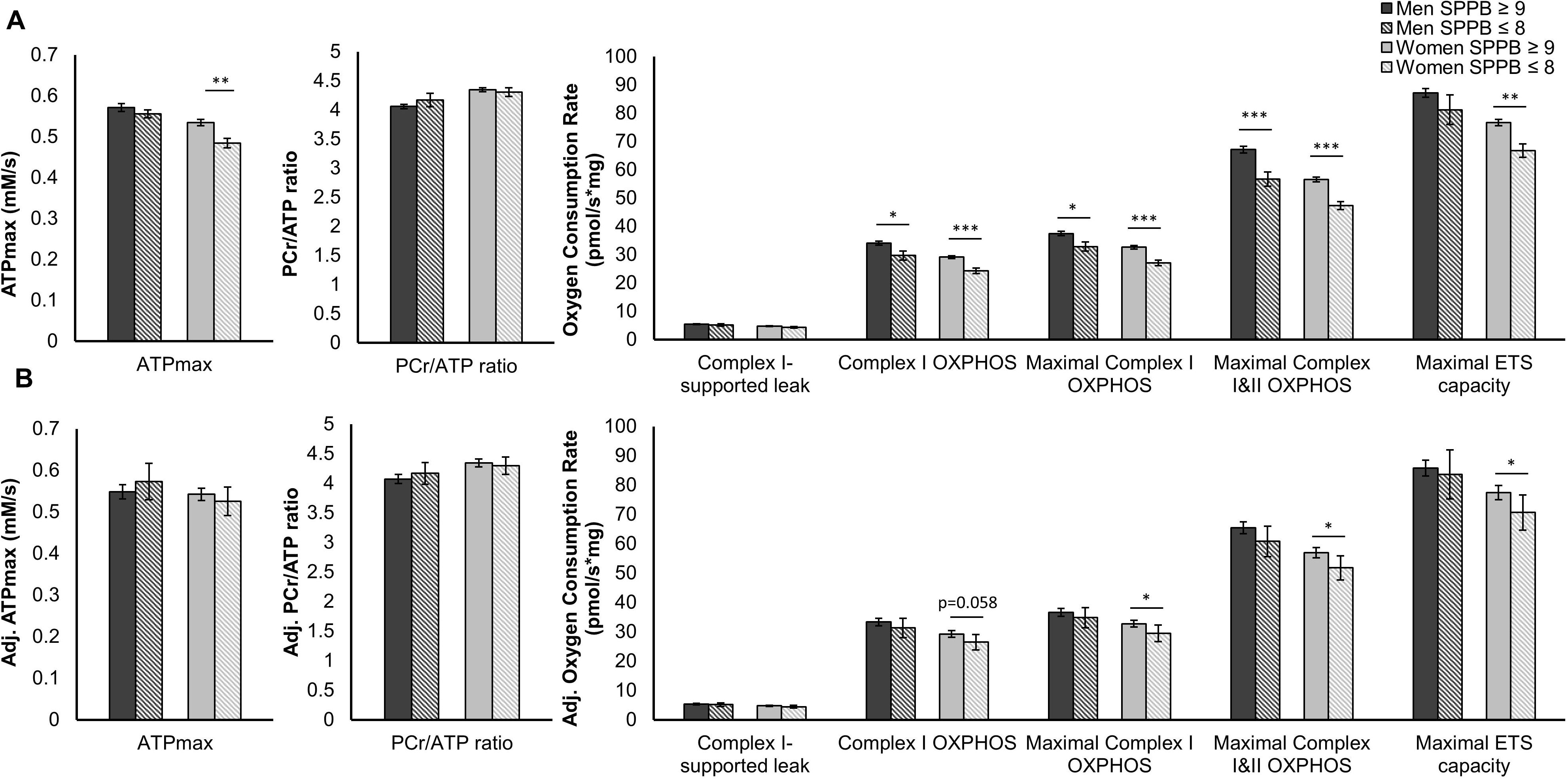
Muscle mitochondrial energetics and mobility impairment. **A**) Unadjusted (+/− SEM) and **B)** covariate (excluding gait speed) adjusted (95% CI) in vivo and ex vivo muscle energetics in men and women with mobility impairment (SPPB ≤ 8) or without mobility impairment (SPPB ≥ 9). ***=p<0.001, **=p<0.01. *=p<0.05.

### Muscle Mitochondrial Energetics and the Odds of Mobility Impairment

To test whether muscle energetics explained sex differences in the prevalence of mobility impairment, we performed mediation modeling (**Figure 3A, B; Supplementary Table 1**). Overall, the odds of women having mobility impairment (SPPB≤8) compared to men was 1.45 (95% CI=0.96, 2.19, p=0.0763) in SOMMA. As the prevalence of mobility impairment in men and women was roughly similar after the age of 80 (**Table 1**), we perform separate models based on age category (70-79, 80+ years of age). As observed in **Figure 3A**, women had significantly greater odds of having mobility impairment compared to men between the ages of 70 and 79 (Model 1: OR=1.83, 95% CI=1.06, 3.15, p=0.0296), which was only slightly mediated by adjustment with age (Model 2: OR_age adjusted_=1.78, 95% CI=1.03, 3.08, p=0.0383). Model 3 of the mediation analysis consisted of a sequential adjustment of Model 2 for a single muscle energetics measure (including technician or site) followed by other covariates, including race, BMI, CHAMPS, and number of chronic health conditions. Each muscle energetics measure mediated the greater odds of mobility impairment in women compared to men. In vivo measures (^31^P MRS-Model 3a, b) were the least mediatory with approximately 25% mediation, while ex vivo measures (HRR-Model 3c-g) mediated the odds by approximately 30 to 70%. These odds were further mediated by the other covariates. In the case of Maximal Complex I & II supported OXPHOS, adjustment mediated the OR to 1.17 (95% CI=0.65, 2.12, p=0.5935), which was further mediated to 0.99 (95% CI=0.52, 1.86, p=0.97) after covariate adjustment. After the age of 80, women no longer had significantly greater odds of mobility impairment (OR=1, 95% CI=0.51, 1.96, p= 0.99), and muscle energetics adjustment did little to change these odds, except in the case of PCr/ATP ratio and Maximal ETS capacity, which increased the odds of women having mobility impairment, but not significantly (**Figure 3B**). Adjusting by covariates alone mediated the sex disparity in mobility impairment to OR=1.21 (0.66, 2.19) in the 70-79 age group and OR=0.96 (0.44, 2.08) in the 80+ age group (**Supplementary Table 1**). To further explore this finding, we assessed these age groups together and fully adjusted (excluding gait speed), for the odds of mobility impairment based on a 1 SD increase in muscle energetics in both men and women (**Figure 3C**). A 1 SD increase in four of the ex vivo muscle energetics measures resulted in significantly lower odds of mobility impairment in women, but not men. These were Complex I OXPHOS (OR=0.62, 95% CI=0.43, 0.90, p=0.0116), Maximal Complex I supported OXPHOS (OR=0.59, 95% CI=0.41, 0.87, p=0.0067), Maximal ETS capacity (OR=0.56, 95% CI=0.37, 0.85, p=0.0062), and Maximal Complex I&II supported OXPHOS (OR=0.57, 95% CI=0.39, 0.84, p=0.0045). To determine if the relationship between muscle energetics measures and mobility impairment differs by sex, we examined the sex/muscle energetics interaction term. The only significant interaction was observed with Maximal ETS capacity (p=0.045), suggesting that higher ETS capacity is associated with a lower likelihood of mobility impairment in women, with little to no association of ETS capacity on the likelihood of mobility impairment in men. To explore the effect of muscle mass and/or physical activity (by actigraphy in place of CHAMPS), additional mediation models were performed (Supplementary Table 1). Adding muscle mass and/or total activity counts by actigraphy (in place of CHAMPS) to the adjusted Model 3 showed similar results to that observed in Figure 3, but with covariates explaining less of the mobility impairment odds. However, accounting for muscle mass and total activity counts individually resulted in disparate results. Total activity counts appear to be a negative confounder, perhaps due to the 1.15 fold higher activity counts in women compared to men, and increased the odds of women having mobility impairment when added to the 70-79 age group covariate adjustment model in place of CHAMPS (OR=2.29, 95% CI=1.23, 4.25, p=0.0087), though muscle energetics still explained much of this disparity (e.g., Maximal Complex I&II supported OXPHOS OR=1.66, 95% CI=0.86, 3.21, p=0.1319). Though initially muscle mass appears to explain all of the gender disparity in mobility impairment in the 70-79 age group (OR=1.01, 95% CI=0.46, 2.24, p=0.9724), possible over-adjustment is apparent in the 80+ age group which previously showed minimal to no differences in the odds of mobility impairment in women pre or post muscle energetics and covariate adjustment, yet accounting for muscle mass shifted the OR to approximately 0.5 for all values. This suggests men were twice as likely to have mobility impairment than women after 80 years of age when accounting for muscle mass. For these reasons, muscle mass and total activity count (in place of CHAMPS) were not included in Figure 3A and B, but made available in Supplementary Table 1.

**Figure 3:**
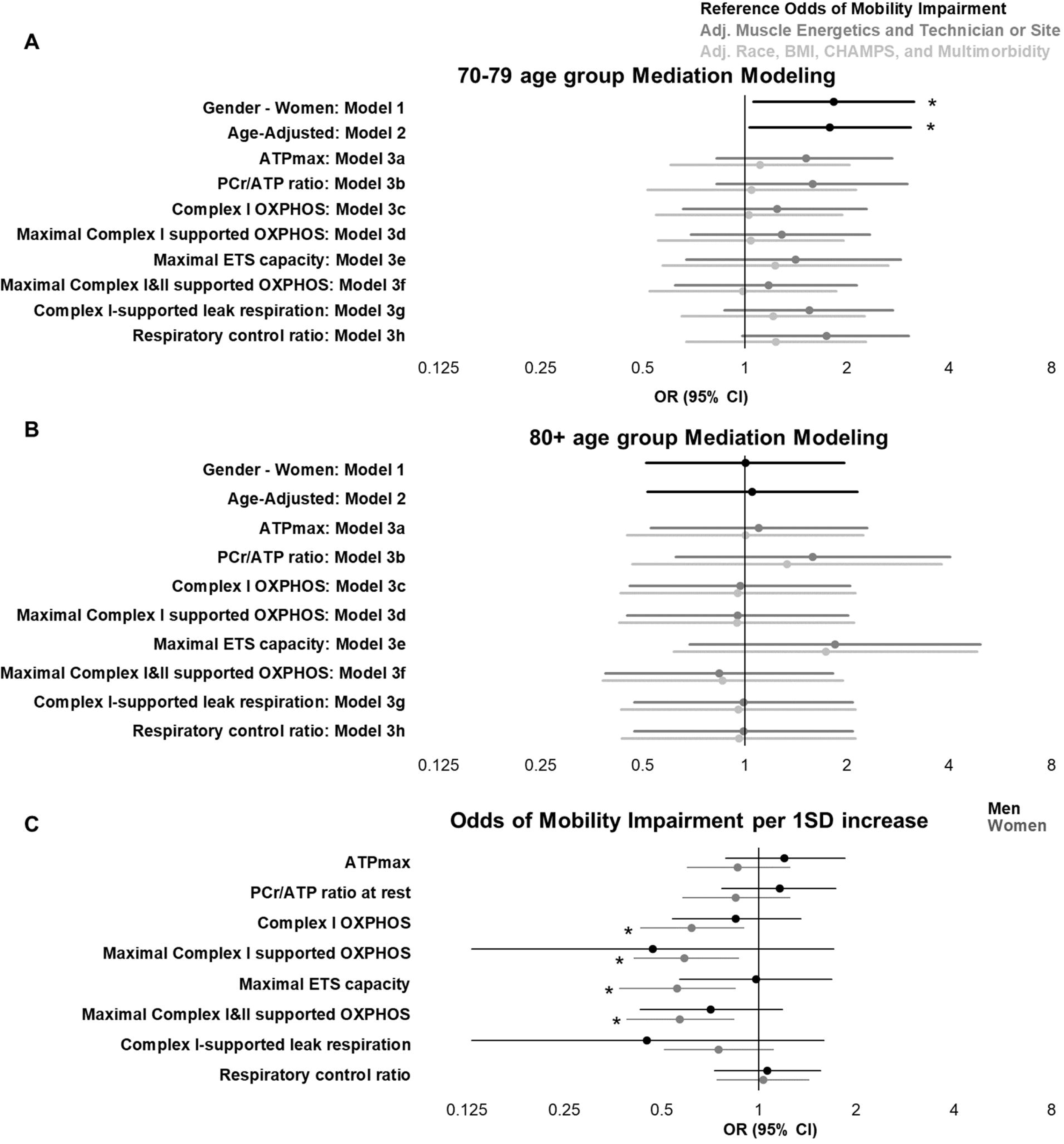
Mediation modeling and the odds of having mobility impairment (SPPB ≤ 8). **A**) Mediation modeling shows the odds ratio (OR and 95% CI) of women having mobility impairment (SPPB≤8) compared to men in the 70-79 age group and the **B)** 80+ age group. Model 2 adjusts for age and Model 3 (a-h) for muscle energetics and technician or site in the case of ATPmax and PCr/ATP ratio at rest, and then adjustment for covariates. **C)** The odds ratio (OR) of mobility impairment for 1SD increase in muscle energetics are expressed as mean and 95% CI. *=p<0.05. Data adjusted for Age, BMI, race, technician, CHAMPS and number of chronic health conditions. The p-values for significant sex and energetics interaction terms are as follows: ATPmax, p=0.168; PCr/ATP ratio at rest, p=0.297; Complex I OXPHOS, p=0.179; Maximal Complex I OXPHOS, p=0.176; Maximal ETS capacity, p=0.045; Maximal Complex I&II OXPHOS, p=0.574; Complex I-supported leak respiration, p=0.248; Respiratory control ratio, p=0.903.

## Discussion

In this analysis of SOMMA participants, women had significantly lower muscle energetics and greater odds of mobility impairment compared to men until the age of 80. Age, BMI, race, physical activity (CHAMPS), gait speed, and number of chronic conditions were unable to explain sex differences in muscle energetics, with the exception of ATPmax. Both men and women with mobility impairment had significantly lower muscle energetics compared to individuals who were not mobility impaired. Covariate adjustment (excluding gait speed) explained the muscle energetics differences due to mobility impairment in men, but not in women, with the exception of ATPmax. Interestingly, muscle mitochondrial energetics were associated with age only in men. We report for the first time that the higher burden of mobility impairment in women before age 80 was largely explained by lower muscle energetics, with age, BMI, race, physical activity (CHAMPS), and number of chronic health conditions providing additional explanatory power. This finding provides a clearer understanding of why women may experience greater early incidence of mobility impairment and suggests muscle mitochondrial energetics as a potential therapeutic target to mitigate the loss of mobility in general and particularly for women. Interventions such as exercise, may be sufficient prevent or reverse mobility impairment based on its known role in improving muscle energetics ^34^. In one study, trained vs untrained older adults had a higher ETS capacity of about 22/pmol/s*mg, enough to reduce the odds of mobility impairment in women by 0.56 as reported here, though larger and longitudinal studies are needed ^23,35^.

We have previously described associations between multiple measures of energetics, such as VO2 peak, ATPmax by ^31^P MRS, and skeletal muscle energetics by HRR, with physical activity and function, such as weekly caloric expenditure (CHAMPS Questionnaire), and leg strength in SOMMA^24^. Here, we report that muscle energetics is also impaired in individuals with mobility impairment, though more so in women than men after accounting for covariates. In SOMMA, lower-extremity mobility impairment was determined using the Short Physical Performance Battery (SPPB). This metric has proven to be an effective screening tool for both men and women for identifying mobility impairments, including major mobility disability and sarcopenia, and does not require correction based on sex as is required for grip strength or lean muscle mass in most sarcopenia diagnostic criteria ^31^. Gait speed, a primary component of the SPPB, is often used to describe mobility impairment and related incidence in older adults ^36,37^. The impact of height on gait speed is one sex-dependent consideration, though this has been shown to associate less in older adults than younger adults ^38^. In SOMMA, there was a significant correlation between height and gait speed (r=0.19, p<0.0001), however, stratified by sex, this correlation only existed in women (r=0.17, p<0.0001) and not men (r=0.04, p=0.48). BMI is also considered a major contributor to sex differences in muscle energetics and function ^15,39–41^. However, while the average BMI was significantly different between men and women, only ATPmax had a significant BMI/sex interaction (p=0.0123).

Participant age was the primary sex-dependent factor affecting muscle energetics and mobility impairment. The effect of age on muscle mitochondrial energetics and mobility has been well described ^2,25^. However, in SOMMA, this negative association appears to be driven by men and across multiple energetics measures. Unexpectedly, no measures of muscle energetics were associated with age in women. In this analysis, the greater prevalence of mobility impairment and the lower muscle energetics in women were most evident before the age of 80, after which mobility impairment prevalence and most energetics measures (except PCr/ATP ratio and Maximal ETS capacity) were similar. This is interesting, as Miotto et al. demonstrated that the muscle in younger men and women averaging 22 years of age had similar maximal respiration ^16^. Additional studies are needed to determine if the age-related decline in muscle energetics and function in women occurs before the age of 70, which was outside the scope of this study. Other evidence suggests this decline occurs during menopause, which is associated with a significant decrease in physical function ^14,42,43^. Indeed, sex hormones have been attributed to many age and sex differences observed in skeletal muscle, such as greater fatty acid oxidation in women, greater mitochondrial ROS generation in men, and morphological differences such as fiber type composition and size, as well as capillary density ^44–46^. However, to our knowledge, sex-based differences in muscle composition and metabolism, including the higher oxidative phosphorylation shown in women in one small respirometry study, have not been replicated in permeabilized fibers and in larger older cohorts ^47–50^. In fact, sex differences in mitochondrial energetics have not been conclusively demonstrated across all tissues let alone across the age spectrum ^17^.

In this analysis, we assessed the relative contribution of study covariates and muscle energetics on the odds of mobility impairment. Between 70-79 years of age, women had 1.78 fold higher odds of mobility impairment than men. Individually adjusting for each energetics measure demonstrated that muscle energetics were capable of explaining one to three quarters of the sex disparity in mobility impairment, with Maximal Complex I&II supported OXPHOS accounting for the greatest attenuation. Further mediation was observed with adjustment to covariates, namely race, BMI, technician or site, CHAMPS, and the number of chronic health conditions. Covariates adjustment alone explained nearly three quarters of the mobility impairment disparity. Together, Maximal Complex I&II supported OXPHOS and covariates fully mediated the sex disparity. This was not observed in individuals older than 80 years of age, where no such mobility impairment disparity existed. Interestingly, adjusting for PCr/ATP ratio and Maximal ETS capacity increased the odds of women having mobility impairment, though not significantly, likely due to the significant sex difference in these two measures in this age group. Including both age groups (70-94) in a fully adjusted model, 1 SD increase in Complex I OXPHOS, Maximal Complex I supported OXPHOS, Maximal ETS capacity, and Maximal Complex I&II supported OXPHOS reduced the odds of mobility impairment. Similarities between Complex I OXPHOS and Maximal Complex I supported OXPHOS in these models are likely due to minor effects of glutamate on total oxygen consumption, which was the only difference between the two measures. When muscle mass and total activity count (as measured by actigraphy) were added to the mediation model, covariates explained less of the mobility impairment disparity, though this is believed to be due to a mixed confounder effect, and each measure was suspected of over-adjustment when accounted for individually. Future studies are needed to assess the contribution of total muscle mass and activity count measures in lower-extremity mobility impairment. Notably, the in vivo measures of ATPmax and PCr/ATP ratio did not explain the mobility impairment sex disparity as markedly as ex vivo measures, and ATPmax was not significantly different in men with mobility impairment but was with women prior to covariate adjustment. Differences between in vivo and ex vivo muscle energetics could be due to technological, methodological, or biological factors, such as differences in the limit of detection and sex difference in adiposity of the thigh, modification of the redox environment ex vivo, and limited substrate availability in vivo. Interestingly, the PCr/ATP ratio was the only muscle energetics measure with significantly higher values in women than men. Since PCr/ATP is a measure of energy homeostasis, this suggests that there may be a small but statistically significant difference in resting energy status in men and women across the ages studied. Further studies are needed to elucidate the specific energetics differences between men and women and their role in mobility impairment, including mitochondrial content, mitochondrial complex expression, fatty acid oxidation, oxidant generation, and oxidative modifications. In addition to the technical limitations stated above, this cross-sectional analysis utilized several self-reported indices, including health history, physical activity, race, and sex. As sex at birth was not determined, no distinction could be made between biological sex and self-identified gender. We acknowledge that both sex and gender may impact this study’s outcomes on physiological and behavioral/cultural levels ^51,52^. Despite these limitations, SOMMA represents one of the largest studies in a deep and comprehensively phenotyped older population with assessments of muscle energetics and physical function.

Muscle mitochondrial energetics are significantly lower in women than men and in individuals with lower-extremity mobility impairment, even after accounting for covariates. Age was a major sex-dependent factor, which associated with muscle energetics only in men. Accounting for muscle energetics attenuated the significantly greater odds of mobility impairment in women in the 70-79 age group. After the age of 80, both muscle mitochondrial energetics and the odds of having mobility impairment were largely similar between men and women. Together, these data suggest the sex disparity in mobility impairment can be largely attributed to sex differences in muscle mitochondrial energetics and its age-related decline. These results provide possible targets for intervention to improve mitochondrial energetics and reduce the burden of mobility impairment and other disabilities, particularly in women.

## Data Availability

All data produced in the present study are available upon reasonable request to the authors.

https://www.sommastudy.com/

## Acknowledgements

We acknowledge all the wonderful staff and investigators of SOMMA including clinical and support staff at Wake Forest University School of Medicine and University of Pittsburgh, as well as the San Francisco Coordinating Center^29^. Author Contributions: P Kramer led the writing team and data meetings and co-drafted the manuscript with A Molina. P Kramer, S Harrison, and P Cawthon led formal analyses. K Duchowny, P Cawthon, A Newman, and P Coen provided the most critical reviews and edits that led to the improvement of the manuscript. G Distefano, S Ramos, E Shankland, and D Marcinek all contributed with either data validation, experimental designs, and/or resources. P Kramer and F Toledo participated in either biopsy collection, processing, and/or experiments. S Cummings, P Coen, A Molina, B Goodpaster, P Cawthon, A Newman, S Kritchevsky, and R Hepple enabled the study with funding acquisition, project administration, and/or conceptualization of the study.

